# Behavioural activation for co- morbid depression in people with non-communicable disease in India: Protocol for a randomised controlled feasibility trial (BEACON)

**DOI:** 10.1101/2022.05.25.22275556

**Authors:** Rayeesa Zainab, Arun Kandasamy, Naseer Ahmad Bhat, Chrishma Violla Dsouza, Hannah Jennings, Cath Jackson, Papiya Mazumdar, Catherine Hewitt, David Ekers, Gitanjali Narayanan, Girish N Rao, Karen Coales, Krishna Prasad Muliyala, Santosh Kumar Chaturvedi, Pratima Murthy, Najma Siddiqi

## Abstract

**Introduction:** The increasing burden of depression and non-communicable disease (NCD) is a global challenge, especially in low- and middle-income countries (LMIC) considering the resource constraints and lack of manpower in these settings. Brief psychological therapies such as behavioural activation (BA), have shown to be effective for the treatment of depression. However, their feasibility and effectiveness for depression in people with NCDs in Indian community setting has not been systematically evaluated. Hence, in this study, we conceptualize to adapt BA into Indian NCD context, thus to improve the management of depression in people with NCD in India.

**Aims:** To (1) adapt BA for the Indian NCD context, (2) test the acceptability, feasibility and implementation of the adapted BA intervention (BEACON intervention package, BIP), and (3) test the feasibility of a randomised controlled trial evaluation of BIP for the treatment of depression compared with enhanced usual care.

**Methods:** Following well-established frameworks for intervention adaptation, we first adapted BA (to fit the linguistic, cultural and resource context) for delivery in India. The intervention was also adapted for potential remote delivery via telephone.

In a randomised controlled trial, we will then test acceptability, feasibility and implementation of the adapted BA intervention (BEACON Intervention package, BIP). We also test if a randomised controlled feasibility trial can be delivered effectively and estimate important parameters (e.g. recruitment and retention rates and completeness of follow up) needed to design a future definitive trial.

Findings will be used to refine procedures for a future definitive trial evaluation of the effectiveness (and cost-effectiveness) of the BIP compared with enhanced usual care for the treatment of depression in NCDs.

**Ethics and dissemination:** The study has received ethics approval by the University of York Health Sciences Research Governance Committee, UK; the Health Ministry Screening Committee, India; and the Ethics Committee (Behavioural Sciences Division), NIMHANS, Bangalore, India.

**Trial Registration:** CTRI/2020/05/025048 [Registered on: 06/05/2020], http://ctri.nic.in

This research was funded by the National Institute for Health Research (NIHR) (17/63/130) using UK aid from the UK Government to support global health research. The views expressed in this publication are those of the author(s) and not necessarily those of the NIHR or the UK government.

## Introduction

Non-communicable diseases (NCDs), also known as chronic diseases, tend to be of long duration and are the result of a combination of genetic, physiological, environmental and behavioural factors. NCDs are the leading cause of mortality in the world, much of which is premature and avoidable (1,2). There is increasing evidence that comorbid mental health conditions such as depression can greatly affect people with physical health conditions (3). For example, people with NCDs are 2-3 times more likely to experience depression (2). The presence of depression worsens NCD symptoms, adversely impacts quality of life, and increases the financial burden for patients and their families (2). Therefore, it is important to recognize and treat depression to improve overall management of NCDs and improve health, quality of life and economic outcomes for these patients,

There is an identified need to integrate mental health care in NCD care settings to help address the high burden of co-morbid depression with NCDs. However, there is a notable gap worldwide between the number of people in need of mental health care and those who receive treatment. This ‘treatment gap’ is especially high in Low- and Middle-Income Countries (LMICs)(4,5). Task shifting is a strategy that has been shown to be successful in helping to address this shortage of specialists, whereby non-specialist health workers are trained to deliver interventions that would usually be provided by specialists (6). This approach is supported by the WHO Mental Health Gap Action Programme (mhGAP)(7).

Behavioural activation (BA) is an evidence-based, simple, structured psychotherapy for the treatment of depression, initially developed in the 1970s. BA aims to break the negative cycle of aversive behaviours and low mood, by increasing engagement in activities that are associated with positive reinforcement. BA therapy is delivered by a trained practitioner, usually over a series of face-to-face therapy sessions, with ‘homework’ tasks that are agreed with the person with depression, to be undertaken between sessions. BA can be delivered through a number of sessions via digital and telephone platforms as well as in-person (2,8,9).

Although there is interest in using BA for depression in people with NCDs, evidence for the effectiveness of this approach has not been clearly established. A systematic review conducted in 2011 found only eight randomised controlled trials of BA in long-term conditions, (with study sample sizes ranging from 20 to 105) (10). Three trials targeted stroke, two were in dementia and one in breast cancer. The remaining two included studies targeting older residents in nursing homes but did not specify the comorbid physical conditions. Since the publication of that review, the INDEPENDENT study, a randomised controlled trial of BA delivered via a multicomponent care model in India has shown effectiveness and cost-effectiveness for treatment of depression in people with diabetes. However, the study intervention was complex and multifaceted, combining collaborative care, decision-support, and population health management. It remains uncertain whether BA is effective for the treatment of depression in NCDs in LMICs.(11)

This study aims to address this evidence gap by (1) adapting BA for the Indian NCD context, (2) testing the acceptability, feasibility and implementation of the adapted BA intervention (BEACON intervention package, BIP), and (3) testing feasibility of randomised controlled trial evaluation of BIP for the treatment of depression compared with enhanced usual care.

Aim 1 has already been completed.

## Methods

### Intervention adaptation

The purpose of this step was to ensure the BA intervention content and delivery were appropriate for the Indian NCD cultural, linguistic and resource context. The initial content was based on treatment manuals and workbooks from previous adaptations of BA for multimorbidity and for delivery by lay counsellors in the UK (12) and India (13). The research team also looked at training packages from the WHO mhGAP (7) relevant to the delivery of BA and formative research exploring the implementation of BA in similar settings in South Asia (14).

Following adaptation frameworks, including for cultural adaptation (15)(16), and taking this information into account, and in discussion with experts within NIMHANS and the wider BEACON team, the materials, plans for delivery and training package were adapted. Revisions to the package were made through expert panel reviews, co-design workshops with NCD counsellors and feedback sessions from NCD patients and caregivers. Major revisions included modification of the workbook as a flipbook with less text and more pictures, simplification of the BA counsellor’s manual, preparation of a separate session log, instead of homework tasks, in between session follow ups were decided. The training package for the BA counsellors was modified in collaboration with a BA expert from UK.

The finalised BIP comprised a BA counsellor’s manual and session log and a flipbook for the patient participants. The intervention could be delivered remotely via telephone, over six sessions lasting approximately 30-40 minutes each. Training of the BA counsellors focused on developing skills in communication, establishing therapeutic relationships, and using the manual and materials to deliver therapy sessions.

### Feasibility trial

#### Objectives

The objectives of the feasibility trial are to:

i. Test the acceptability, feasibility and implementation of the adapted BA intervention (BEACON Intervention package, BIP)
ii. Test if a randomised controlled feasibility trial can be delivered effectively and to estimate important parameters needed to design a future definitive trial

These will be achieved through answering the following research questions (RQs):

1. What are the recruitment and retention rates for participants in the trial?
2. What is the feasibility and acceptability of proposed randomisation and data collection procedures?
3. To what degree can the BIP be implemented as planned?
4. What is the acceptability of the BIP from the perspective of study participants and BA counsellors?

#### Design

A randomised controlled feasibility trial with nested process and economic evaluations will be conducted over 12 months. The GANNT chart is presented in Appendix 1.

The two trial arms are:

- Intervention arm: BIP
- Control arm: Enhanced usual care

#### Setting

In 2010 the Ministry of Health and Family Welfare, Government of India launched the National Programme for Prevention and Control of Cancer, Diabetes, Cardiovascular diseases and Stroke (NPCDCS). This includes the prevention and management of NCDs at district and sub-district level facilities. These facilities provide services for screening, treatment or referral of patients with NCD, but do not, at present, include the integrated detection and management of mental disorders. They do not yet cover all districts in the country. (17)

The setting for the BEACON study is a district (Kolar, Karnataka, South India) where it will be possible to explore the integration of mental and physical health care, based on several pragmatic considerations, including the availability of NCD services that are part of the government health system, and engagement of key stakeholders. Selection of healthcare facilities will be based on i) provision of NCD services, ii) availability of healthcare staff such as NCD counsellors and nurses to identify the patient participants for the study (by administering the Patient Health Questionnaire (PHQ-2)(18), a screening tool to identify those who need further assessment for depression) iii) engagement and support of senior managers, and iv) a sufficient number of patients with NCD attending the facility. Facilities will be part of government-supported health services and be within a reasonable travelling distance of the research team’s base to make the study feasible.

Since most NCD services are limited to providing care for cardiovascular and chronic lung diseases and diabetes (excluding cancer), the study will focus on these disorders.

#### Eligibility criteria for trial participants

*Inclusion criteria:*

- Aged 18 years or over
- Diagnosed with cardiovascular disease, chronic respiratory disease or diabetes (type 1 or 2)
- Current diagnosis of depression (confirmed with PHQ-9(19) (score of ≥ 10).
- Willing to participate and able to attend therapy sessions in person or by telephone.
- Other mental or physical illness comorbidities will not be a reason to exclude unless the patient is judged to be too unwell to participate.

*Exclusion criteria:*

- Already receiving psychotherapy for depression. Being on antidepressant medication will not however be a reason to exclude.
- Lacking capacity to provide informed consent.
- Unable to take part in therapy because of cognitive impairment, or severity of mental or physical illness.

#### Intervention arm

Participants in the intervention arm will receive the BIP, delivered by trained non-mental health specialists (BA counsellors) who will be specifically recruited for the study. BA counsellors will be supervised by a mental health specialist.

The BIP will be delivered over six sessions lasting 30-40-minutes over a period of 6 to 12 weeks, with a minimum of one week between sessions. All sessions will be delivered remotely, by telephone.

The components of BIP include structured sessions where participants, with the BA counsellors, plan and identify three types of activities (routine and necessary, social, pleasurable) to complete and will monitor their mood accordingly. They will have a flipbook for guidance. The BA counsellor will follow a manual and maintain the session log of the participants.

#### Control arm

Participants in the control arm will receive an enhanced usual care leaflet describing depression and its treatment, and signposting, including providing contact details (addresses and telephone numbers) to help access usual care for depression. It is recognised that depression management is not standardised across services and is often not available at all.

#### Outcomes

Outcomes for the feasibility trial relate to i) acceptability and feasibility of recruitment and data collection, and ii) acceptability, feasibility and implementation of the BIP.

*Outcomes relating to feasibility and acceptability of recruitment and data collection:*

- Rates of recruitment into the trial and retention at 3 months (RQ1)
- Proportion of participants recruited who are randomised (RQ2)
- Completeness of data collection (% of planned measures completed) (RQ2) at:
  - <u>Baseline:</u> Demographic data, Depression symptoms (Patient Health Questionnaire-9 (PHQ-9) (19), Depression anxiety and stress scale (DASS) (20), Anxiety (GAD-7) (21), Health-related quality of life (Euroqol (EQ-5D-5L) (22), Measures of potential mediators: e.g. knowledge, intention, beliefs about consequences, Premium Accredited Activation Scale (PAAS) (23), Adverse events
  - <u>3 months:</u> Depression symptoms PHQ-9 (Likely to be the primary outcome for the full trial), GAD-7 (21), EQ-5D-5L (22), (PAAS) (23), Adverse events

Outcomes relating to (ii) acceptability, feasibility and implementation of the BIP are described below in the process evaluation.

#### Trial sample size

The sample size is set to enable reliable estimation of recruitment and retention rates, and to adequately explore the feasibility of data collection and intervention delivery in this feasibility trial. 56 participants will be recruited into the study. This should be adequate to estimate a participation rate of 20% and a follow-up rate of 80% to within a 95% confidence interval of ±5% and ±10% respectively.

#### Trial recruitment and consent procedures

The BEACON study flow chart is presented in Appendix 2.

Participating sites will be asked to implement depression screening by NCD healthcare staff such as NCD counsellors and nurses using the Patient Health Questionnaire-2 (PHQ-2) (18) as part of routine clinical practice. The PHQ-2 is a simple screening tool, with minimal training and time requirements. Training will be provided by the study team, and staff asked to use the PHQ-2 with every patient attending the NCD service.

Staff (NCD counsellors and nurses) will be asked to give information about the study to patients who screen positive for PHQ-2 (score ≥ 3). Those who indicate they are interested in participating in the feasibility trial will be approached by research assistants for the next level screening, by administering the PHQ-9 (19).

Recruitment of the willing participants by research assistants will be done either face to face or remotely by telephone, mobile phone, internet or video calling as is convenient for the participant. Accordingly, written informed consent or verbal informed consent (in case of remote recruitment) will be obtained prior to administering the PHQ-9 along with consent for participation in the feasibility trial. Those screening positive for depression on PHQ-9 (score ≥ 10) will be invited to participate in the feasibility trial, and provided detailed information about the purpose of the study and trial procedures either face to face or remotely. And final consent for participating in the trial will be obtained. Also, suicide risk assessment will be conducted at this point.

#### Randomisation

Once consent has been secured, eligible participants will complete a baseline questionnaire and will then be randomised (BIP or enhanced usual care) by the trial manager/Research Fellow who will be based at the central office. Participants will be allocated in a 1:1 ratio using simple randomisation without stratification. Treatment allocation will be concealed from the study team at the point of recruitment using an automated computer data entry system, administered remotely by York Trials Unit and using a computer-generated randomisation sequence generated by an independent statistician (using Stata 16 or later).

#### Data collection

Baseline and 3-month follow-up data will be collected by research assistants (See Table 1: SPIRIT schedule of enrolment, interventions and assessments). Quantitative data will be collected with predesigned and tested data collection forms, using tablets to improve efficiency and to minimise risk of errors in data entry. Data will be anonymised replacing identifiable personal data with unique study participant identification numbers, with the key only available to the local research team PI and manager. Data will be secured and transferred safely, in line with University of York data management policies and procedures.

**Table 1:**
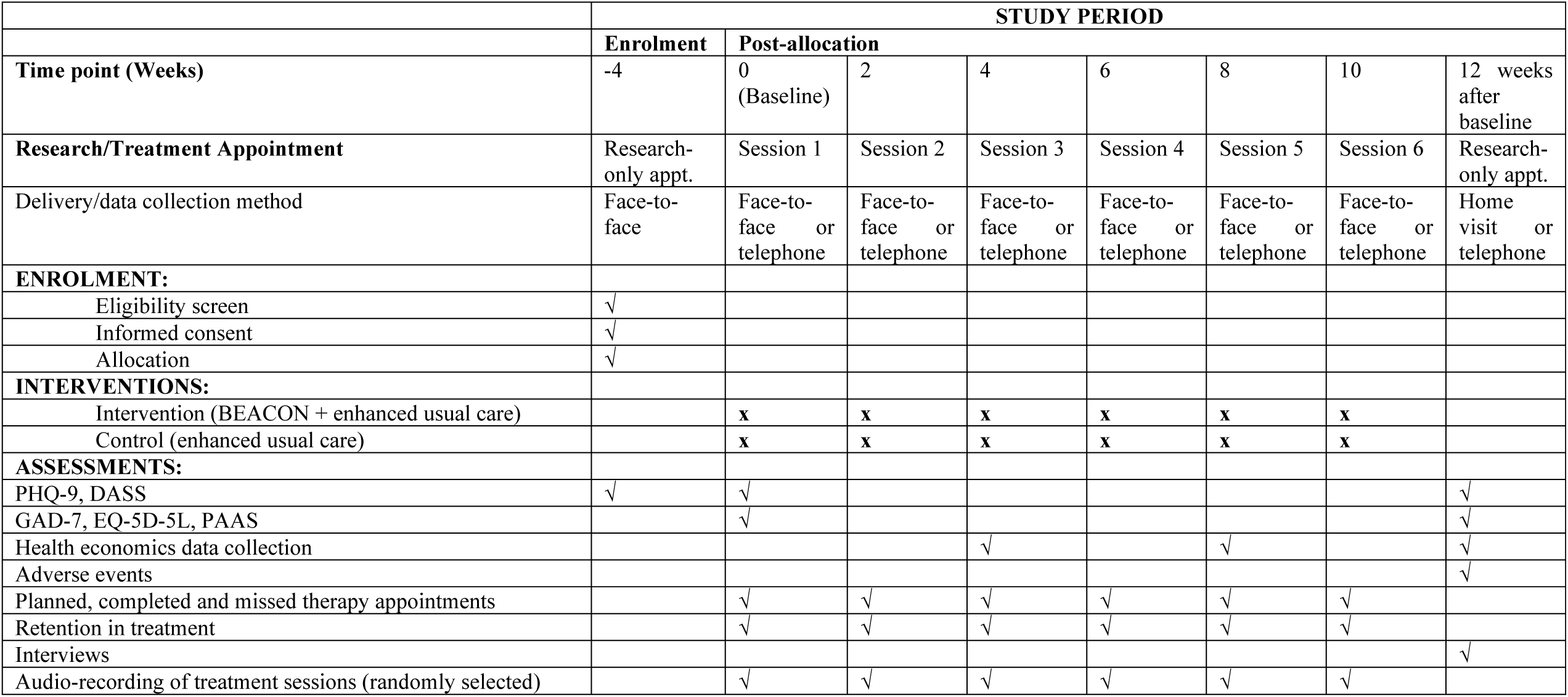
SPIRIT schedule of enrolment, interventions and assessments:

#### Quantitative data analysis

Analyses will be conducted using Stata software. The number of participants who were screened, randomised, treated, and followed up for the primary outcome (PHQ-9) (19) will be reported in a CONSORT-style flow chart (24). The number of study completers, by trial arm, will be calculated as a proportion of the number randomised at baseline.

The proportion of baseline and outcome measures completed out of those planned, and present these by treatment allocation arm will also be calculated.

#### Process evaluation

The three key functions of a process evaluation (mechanisms of impact, context, implementation) will underpin our approach for the feasibility trial (25). Qualitative inquiry will be used in combination with quantitative data to gain a more complete picture.

##### Survey (implementation, mechanisms of impact)

To capture the views and experiences of participants, all participants will complete a brief quantitative survey at 3-month follow up. Participants in the intervention arm will be asked about their engagement with, and acceptability of BIP, views on remote delivery as well as the perceived impact on their depression symptoms. Control arm participants will answer these questions in relation to the enhanced usual care leaflet. Participants in both trial arms will also complete some questions to assess the acceptability of trial processes.

Potential mediators of intervention impact will be measured at baseline and 3-months: intentions, beliefs about capabilities, beliefs about outcomes. These have been selected to reflect the hypothesised mechanisms of action of BA (26).

##### Interviews (implementation, mechanisms of impact and context)

A sub-sample of 16 participants will be interviewed. We will purposively sample to ensure a mix of participants (intervention/control arms, men/women, under 60 years/ 60+ years of age, completed/dropped out of BIP). The interviews will seek in-depth feedback on the topics explored in the brief survey as well as explore individual and context barriers and drivers to performing the three types of activities (routine and necessary, social, pleasurable).

Once delivery of the BIP is completed, a group interview will be conducted with the BA counsellors to explore their experiences, including the acceptability of the interventions and the barriers and drivers to delivery including contextual factors such as remote delivery.

All interviews will be conducted in the local language, face-to-face using topic guides and digitally audio recorded. A hermeneutics approach, which encourages participants to discuss features of the intervention to elicit data on their experience and evaluation of its delivery/receipt will be used with trial participants, family members and healthcare staff (27).

Trial participants will provide written informed consent for these interviews within their trial consent.

##### Fidelity index (implementation)

Fidelity to delivering BIP will be assessed using a fidelity index. This index consists of two sub-indices: The Adherence Index that assesses adherence to delivering the content of the six BIP sessions; and the Interaction Index, that assesses the level of counsellor-participant with which the intervention was delivered. Both indices are scored on a three-point Likert scale (0=not implemented, 1=partially implemented and 2=fully implemented).

All BIP sessions will be audio-recorded. We will then perform the fidelity checks by reviewing the audio-recording of a sub-sample (n=18) of sessions. These sessions will be purposively selected to include a mix of sessions 1-6, delivered by all BA counsellors early and late in the feasibility trial.

##### Data analysis

The quantitative data from the fidelity index and questionnaire will be analysed using descriptive statistics, including means and standard deviation for continuous variables, and absolute and relative frequencies for categorical variables.

Interviews will be transcribed verbatim, translated into English and analysed using the Framework Approach (28). which is particularly useful for understanding and improving programmes/policies and when multiple researchers are working with the data (29). Excel software will aid data handling.

Integration of interview findings with respective questionnaire data will be done using a ‘triangulation protocol’ (30).

#### Economic evaluation

The feasibility of undertaking a full cost-effectiveness analysis will be assessed. This will primarily investigate the acceptability and resource use associated with capturing data for a cost-effectiveness evaluation in the full trial.

The team will capture costs associated with:

i. Depression in NCD
ii. Delivery/receipt of the BIP. Costs will include the time to deliver the BIP and the cost of the materials used.
iii. The costs of training the BA counsellors to deliver the BIP will be calculated. Training requires staff time of the trainer plus staff travel cost. Staff time is based on the salary of the trainer and allocated on a cost per minute basis plus costs of materials.

Diaries to capture resource use, to be completed by patient and BA counsellors, will be developed by the research team. Participants will be asked to complete these on a monthly basis for 3 months.

A proforma to capture costs of delivering the BIP incurred by the host organisation will be completed by service managers, including staff salaries and time, and costs of estates and materials. An average cost per case per trial arm for treatment and the control group will be calculated.

Health service utilisation will be assessed by asking pre-tested questions on contacts with doctors/nurses, hospital admissions, pharmacy visits and drug use etc.

Quality of life will be assessed using a quality of life questionnaire (Euroqol (EQ-5D-5L) at baseline and 3 months. This information can be used as a quantitative measure of health outcomes as judged by the individual respondents. A higher score represents a better-perceived health state. The data will be checked for completeness.

#### Data management

The following data will be collected from feasibility trial participants (at baseline and 3-month follow-up): demographic data, validated instrument measurements of mental and physical health. The process evaluation of the feasibility trial will collect data from interviews with trial participants and BA counsellors, a sample of audio-recorded BIP sessions and drop-out data. Health economic data will be collected to provide evidence of the costs of providing the BIP in the host institutions.

All study data will be stored in accordance with GDPR and the University of York data management policies. Electronic data will be password protected and stored on secure servers at the country partner institutions and at the University of York.

Unique study identification (ID) numbers, allocated by the research team, will be used to anonymise data on electronic documents containing data from the trial and interviews. Paper-based documents, for example consent forms, hard-copy of transcripts as well as any research participant personal data collected will be stored in locked filing cabinets at the partner institutions.

Study documents (paper and electronic) at the research sites will be retained in a secure (locked when not in use) location during and after the study finishes. Interview recordings will be transferred from the digital recorder to a secure server as soon as possible after the interview has taken place. Once the transferred recording has been checked to ensure audibility, the original will be deleted from the digital recorder. All essential documents, including source documents (e.g. transcripts) will be retained for a minimum period of 10 years after study completion, in accordance with the University of York Research Data Management Policy. This is deemed to be sufficient time for any queries arising from the findings to be answered, e.g. queries arising from the publication of findings. They will be retained in secure locations (locked when not in use) at the University of York and at the partner Institutions.

The Department of Health Sciences, University of York has a back-up procedure approved by auditors for disaster recovery. There will be a separate archive of electronic data performed at the end of the study, to safeguard the data, and in accordance with regulatory requirements. The access, use and storage of sensitive or confidential data will be conducted in accordance with the University of York Data Security Policy and Handling Sensitive Data Guidance. All sensitive or confidential data utilised for BEACON will be encrypted.

#### Data monitoring

Data will be monitored for quality and completeness by the delegated researchers at the study site, followed by a second check by researchers at the University of York using verification, validation and checking processes. Missing data will be pursued until study’s end unless it causes any distress to the participant contacted. Data will be reported to the Health Ministry Screening Committee and Ethics Committee as required.

#### Ethical considerations

Working with a potentially vulnerable group of participants has many ethical implications. We have addressed them in our ethics application (see below). The main adverse event anticipated in this study both during the recruitment phase and during delivery of the BIP is self-harm and suicide risk. Therefore, standard operating procedures to address these adverse events will be adapted by the study team.

Ethics and other relevant approvals on the protocol have been secured t from the University of York Health Sciences Research Governance Committee (Approval number: HSRGC/2020/418/B, 27^th^ November 2020), relevant national and institutional ethics committees (i.e., Health Ministry Screening Committee, India – Approval number: 2020-9330 on 12^th^ June 2020); Ethics Committee (Behavioural Sciences Division), NIMHANS (Approval number: NIMHANS/EC (BEH. SC.DIV.) 22”d MEETING/2O I 9); The trial is also registered under the Clinical Trial Registry of India (Approval number: CTRI/2020/05/025048, on 6^th^ May 2020).

Approvals for necessary amendments adjusting to the COVID -19 pandemic situation have been provided by the institutional ethics committee, (Behavioural Sciences Division), NIMHANS

#### Community Engagement

A Community Advisory Panel will provide community engagement and involvement for the duration of the study. This panel comprises patient and family representatives and community and voluntary sector advocates, interested in improving the care of common mental disorders and NCD. The panel has already given feedback on:

∘ The accessibility and feasibility for patients of the proposed BIP intervention, including remote delivery.
∘ Delivery of BA counselling by the NCD Counsellors.
∘ The acceptability of proposed methods for the feasibility trial.

The feedback and suggestions were incorporated in the intervention development and delivery plans of the study. The panel will continue to provide feedback, providing an important perspective on delivery of the study and its findings.

## Discussion

Effective treatment of depression in NCDs offers the potential to improve mental and physical outcomes for this population, and additionally to maximise efficient use of healthcare resources. If shown to be effective, the approach of integrating treatment for a mental disorder within NCD care, will also provide the opportunity to harness physical health resources to reduce the large mental health treatment gap, and to reduce stigma for people with mental illness.

If the feasibility trial demonstrates that the BIP is feasible and potentially beneficial, we will be able to make further adaptations to the BIP and test the intervention in a full trial.

## Data Availability

All data produced in the present study are available upon reasonable request to the authors

## Funding

This research is funded by the National Institute for Health Research (NIHR) (17/63/130) using UK aid from the UK Government to support global health research. The views expressed in this publication are those of the author(s) and not necessarily those of the NIHR or the UK government. The funders have no role study design, data collection and analysis, decision to publish, or preparation of the manuscript.

## Authors Contribution

<u>Conceptualization</u>

Najma Siddiqi

<u>Contextual Information</u>: Arun Kandasamy, Pratima Murthy

<u>Funding Acquisition</u>: Najma Siddiqui

<u>Methodology</u>:

<u>Phase I</u>: David Ekers, Arun Kandasamy, Pratima Murthy, Gitanjali Narayanan, Girish N Rao, Papiya Mazumdar, Karen Coales

<u>Phase II</u>: Najma Siddiqi, Arun Kandasamy, Pratima Murthy, Catherine Hewitt, Catherine Jackson, Hannah Jennings

<u>Manuscript writing</u>: Rayeesa Zainab, Arun Kandasamy, Hannah Jennings, Catherine Jackson

<u>Review and Editing</u>: Chrishma Violla Dsouza, Najma Siddiqi, Krishna Prasad M, Catherine Jackson, Hannah Jennings, Papiya Mazumdar, Karen Coales, Naseer Ahmad Bhat, Pratima Murthy, Gitanjali Narayanan, Girish N Rao, Catherine Hewitt, Santhosh Kumar Chaturvedi, David Ekers, Rayeesa Zainab, Arun Kandasamy

## Acknowledgements

We would like to thank the members of the Community Advisory Panel and the expert panel for their inputs and suggestions for the research. We also would like to thank the study participants for their participation in the study.

## Conflict of interest: None

## Appendices

**Appendix 1:**
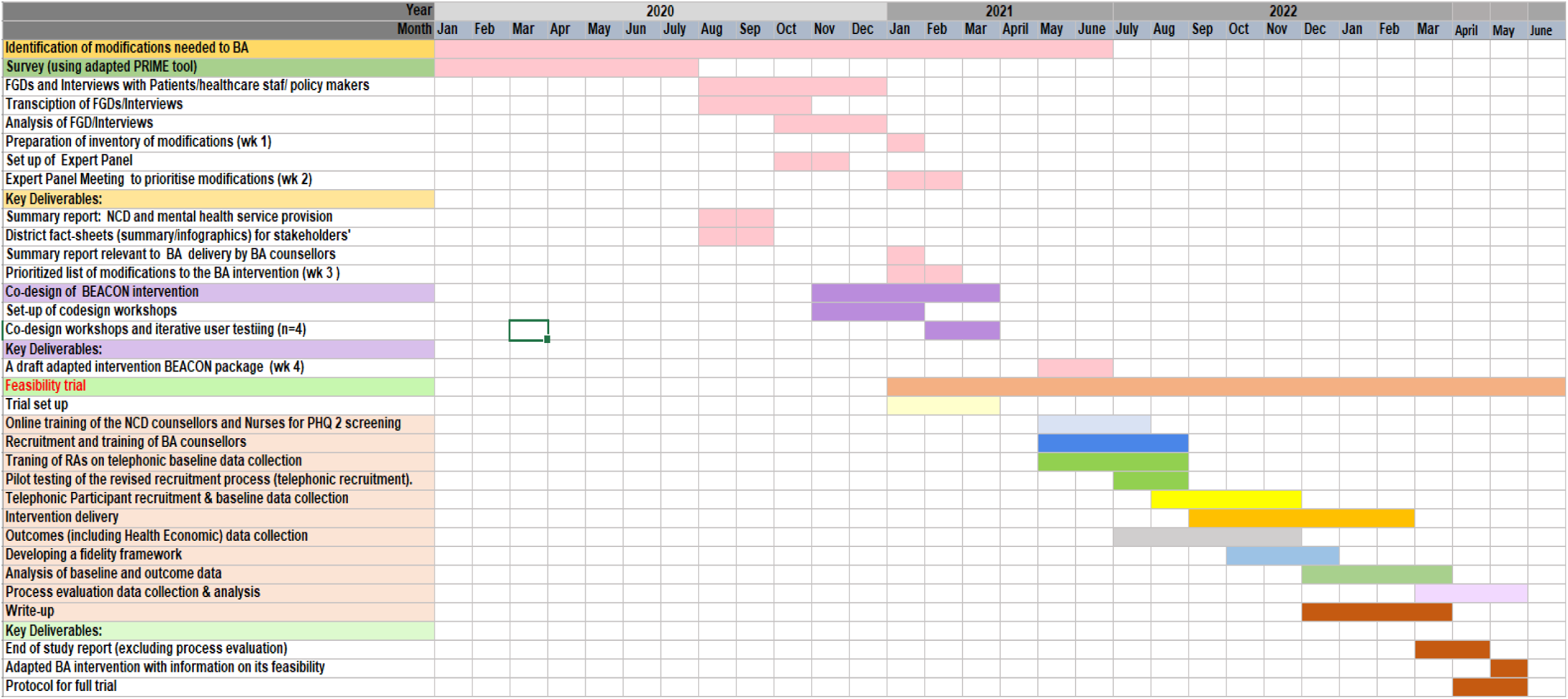
Gantt chart

**Appendix 2:**
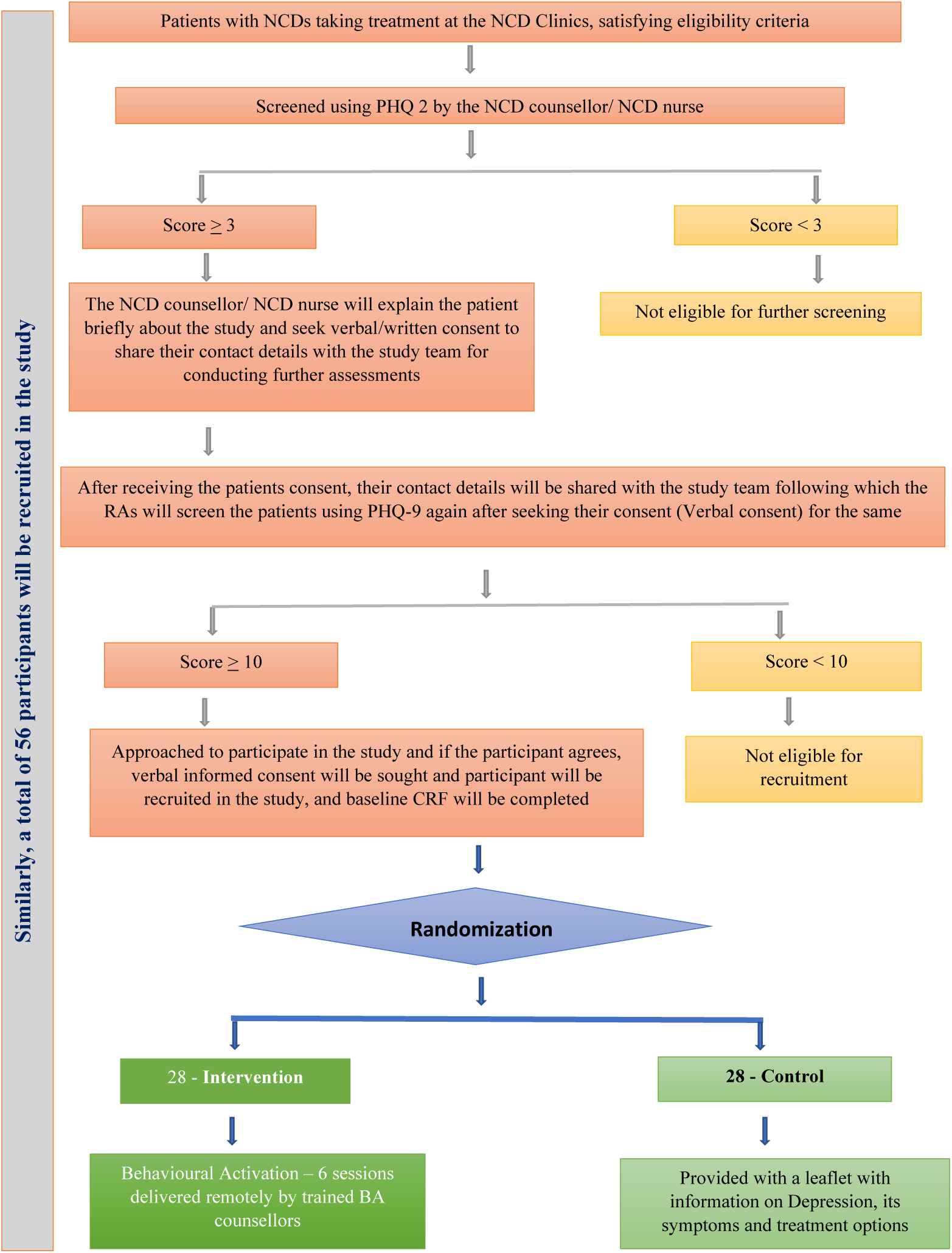
BEACON study flow chart

## Notes

### Competing Interest Statement

The authors have declared no competing interest.

### Clinical Trial

CTRI/2020/05/025048

### Funding Statement

This study was funded by the National Institute for Health Research (NIHR) (17/63/130) using UK aid from the UK Government to support global health research. The views expressed in this publication are those of the author(s) and not necessarily those of the NIHR or the UK government.

### Author Declarations

The University of York Health Sciences Research Governance Committee, UK; the Health Ministry Screening Committee, India; and the Ethics Committee (Behavioural Sciences Division), NIMHANS, Bangalore, India gave ethical approval for this work.

## References

1. World Health Organization. yNoncommunicable diseases. News room Fact sheet. 2021. p. 1.

2. Ngo VK, Rubinstein A, Ganju V, Kanellis P, Loza N, Rabadan-Diehl C, et al. Grand Challenges: Integrating Mental Health Care into the Non-Communicable Disease Agenda. PLoS Med. 2013;10(5):1–5.

3. Naylor C, Parsonage M, McDaid D, Knapp M, Fossey M, A G. Long-Term Conditions and Mental Health: The Cost of Co-Morbidities. 2012.

4. Chisholm D, Flisher AJ, Lund C, Patel V, Saxena S, Thornicroft G, et al. Scale up services for mental disorders: a call for action. Lancet (London, England). 2007 Oct;370(9594):1241–52.

5. Wang PS, Aguilar-Gaxiola S, Alonso J, Angermeyer MC, Borges G, Bromet EJ, et al. Use of mental health services for anxiety, mood, and substance disorders in 17 countries in the WHO world mental health surveys. Lancet (London, England). 2007 Sep;370(9590):841–50.

6. Kakuma R, Minas H, van Ginneken N, Dal Poz MR, Desiraju K, Morris JE, et al. Human resources for mental health care: current situation and strategies for action. Lancet (London, England). 2011 Nov;378(9803):1654–63.

7. World Health Organization. No Title. mhGAP : Mental Health Gap Action Programme: scaling up care for mental, neurological and substance use disorders. 2008. p. 36 p.

8. Ekers D, Webster L, Van Straten A, Cuijpers P, Richards D, Gilbody S. Behavioural activation for depression; an update of meta-analysis of effectiveness and sub group analysis. PLoS One. 2014;9(6):e100100.

9. Kanter JW, Puspitasari AJ, Santos MM, Nagy GA. Behavioural activation: history, evidence and promise. Vol. 200, The British journal of psychiatry : the journal of mental science. England; 2012. p. 361–3.

10. Harris SJ. Behavioural Activation for Depression in Long Term Conditions: Developing a Behavioural Activation Guided Self-Help Intervention for Depression in Dementia. University of Exeter; 2017.

11. Ali MK, Chwastiak L, Poongothai S, Emmert-Fees Kmf, Patel SA, Anjana RM, et al. Effect of a Collaborative Care Model on Depressive Symptoms and Glycated Hemoglobin, Blood Pressure, and Serum Cholesterol among Patients with Depression and Diabetes in India: The INDEPENDENT Randomized Clinical Trial. JAMA -J Am Med Assoc. 2020;324(7):651–62.

12. Gilbody S, Lewis H, Adamson J, Atherton K, Bailey D, Birtwistle J, et al. Effect of Collaborative Care vs Usual Care on Depressive Symptoms in Older Adults With Subthreshold Depression: The CASPER Randomized Clinical Trial. JAMA. 2017 Feb;317(7):728–37.

13. Patel V, Weobong B, Weiss HA, Anand A, Bhat B, Katti B, et al. The Healthy Activity Program (HAP), a lay counsellor-delivered brief psychological treatment for severe depression, in primary care in India: a randomised controlled trial. Lancet (London, England). 2017 Jan;389(10065):176–85.

14. Wright J, Mazumdar P, Barua D, Lina S, Bibi H, Kanwal A, et al. Integrating depression care within NCD provision in Bangladesh and Pakistan: a qualitative study. Int J Ment Health Syst. 2020;14(1):63.

15. Bartholomew LK, Parcel GS, Kok G. Intervention Mapping: A Process for Developing Theory-and Evidence-Based Health Education Programs. Heal Educ Behav. 1998;25(5):545–63.

16. Bernal G. Cultural adaptation : Tools for evidence based practice with diverse populations. 2014;(May 2012).

17. Ministry of Health & Family Welfare Government of India. NATIONAL PROGRAMME FOR PREVENTION & CONTROL OF CANCER, DIABETES, CARDIOVASCULAR DISEASES & STROKE (NPCDCS). Home » NHM Components » Non Communicable Disease; Diabetes; Cardiovascular Diseases & stroke (NPCDCS). p. 1.

18. Kroenke K, Spitzer RL, Williams JBW. The Patient Health Questionnaire-2: Validity of a Two-Item Depression Screener. Med Care. 2003;41:1284–92.

19. Kroenke K, Spitzer RL, Williams JB. The PHQ-9: validity of a brief depression severity measure. J Gen Intern Med. 2001 Sep;16(9):606–13.

20. Sharma MK, Hallford DJ, Anand N. Confirmatory factor analysis of the Depression, Anxiety, and Stress Scale among Indian adults. Indian J Psychiatry. 2020/07/27. 2020;62(4):379–83.

21. Spitzer RL, Kroenke K, Williams JBW, Löwe B. A brief measure for assessing generalized anxiety disorder: the GAD-7. Arch Intern Med. 2006 May;166(10):1092–7.

22. EuroQol--a new facility for the measurement of health-related quality of life. Health Policy. 1990 Dec;16(3):199–208.

23. Patel AR, Weobong B, Patel VH, Singla DR. Psychological treatments for depression among women experiencing intimate partner violence: findings from a randomized controlled trial for behavioral activation in Goa, India. Arch Womens Ment Health. 2019 Dec;22(6):779–89.

24. Boutron I, Moher D, Altman DG, Schulz KF, Ravaud P. Methods and processes of the CONSORT Group: example of an extension for trials assessing nonpharmacologic treatments. Ann Intern Med. 2008 Feb;148(4):W60–6.

25. Moore GF, Audrey S, Barker M, Bond L, Bonell C, Hardeman W, et al. Process evaluation of complex interventions: Medical Research Council guidance. BMJ Br Med J. 2015 Mar;350:h1258.

26. Carey RN, Connell LE, Johnston M, Rothman AJ, de Bruin M, Kelly MP, et al. Behavior Change Techniques and Their Mechanisms of Action: A Synthesis of Links Described in Published Intervention Literature. Ann Behav Med. 2019 Jul;53(8):693–707.

27. Thirsk LM, Clark AM. Using Qualitative Research for Complex Interventions: The Contributions of Hermeneutics. Int J Qual Methods. 2017 Aug;16(1):1609406917721068.

28. Liz Spencer, Jane Ritchie WO. QUALITATIVE RESEARCH PRACTICE A Guide for Social Science Students and Researchers. S JEREADJEL, editor. london: SAGE Publications; 2003. 199–218 p.

29. Gale NK, Heath G, Cameron E, Rashid S, Redwood S. Using the framework method for the analysis of qualitative data in multi-disciplinary health research. BMC Med Res Methodol. 2013;13(1):117.

30. O’Cathain A. A Practical Guide to Using Qualitative Research with Randomized Controlled Trials. Oxford University Press;

